# Assessment of Genetic Susceptibility to Multiple Primary Cancers through Whole-Exome Sequencing in Two Large Multi-Ancestry Studies

**DOI:** 10.1101/2022.02.11.22270688

**Authors:** Taylor B. Cavazos, Linda Kachuri, Rebecca E. Graff, Jovia L. Nierenberg, Khanh K. Thai, Stacey Alexeeff, Stephen Van Den Eeden, Douglas A. Corley, Lawrence H. Kushi, Regeneron Genetics Center, Thomas J. Hoffmann, Elad Ziv, Laurie Habel, Eric Jorgenson, Lori C. Sakoda, John S. Witte

**Affiliations:** Biological and Medical Informatics, University of California San Francisco, San Francisco, CA 94158; Department of Epidemiology and Biostatistics, University of California San Francisco, San Francisco, CA 94158; Division of Research, Kaiser Permanente Northern California, Oakland, CA 94612; Regeneron Genetics Center, Tarrytown, New York 10591; Department of Medicine, University of California San Francisco, San Francisco, CA 94158; Department of Health Systems Science, Kaiser Permanente Bernard J. Tyson School of Medicine, Pasadena, CA 91101; Department of Epidemiology and Population Health, Stanford University, Stanford, CA 94305; Department of Biomedical Data Science, Stanford University, Stanford, CA 94305

## Abstract

Up to one of every six individuals diagnosed with one cancer will be diagnosed with a second primary cancer in their lifetime. Genetic factors contributing to the development of multiple primary cancers, beyond known cancer syndromes, have been underexplored. To characterize genetic susceptibility to multiple cancers, we conducted a pan-cancer, whole-exome sequencing study of individuals drawn from two large prospective cohorts (6,429 cases, 165,853 controls). We created two groupings of individuals diagnosed with multiple primary cancers: 1) an overall combined set with at least two cancers across any of 36 organ sites; and 2) cancer-specific sets defined by an index cancer at one of 16 organ sites with at least 50 cases from each study population. We then investigated whether variants identified from exome sequencing were associated with these sets of multiple cancer cases in comparison to individuals with one and, separately, no cancers. We identified 22 variant-phenotype associations, 10 of which have not been previously discovered and were significantly overrepresented among individuals with multiple cancers, compared to those with a single cancer. Overall, we describe variants and genes that may play a fundamental role in the development of multiple primary cancers and improve our understanding of shared mechanisms underlying carcinogenesis. Further investigation of these findings may lead to new screening strategies for individuals at risk for multiple primary cancers.

## INTRODUCTION

The substantial global burden of cancer coupled with increasing survival due to improved screening, surveillance, and treatments has yielded a growing number of cancer survivors who are at risk of developing a second primary cancer in their lifetime^1,2^. The prevalence of multiple primary cancers globally is estimated to be between 2 and 17%, with the wide range likely due to differences in cancer registration practices, case definitions, population characteristics, and follow-up times^1,2^. Cancer predisposition syndromes, such as Li-Fraumeni, Lynch, and hereditary breast and ovarian cancer, are known to increase the risk of multiple primary cancers; however, less than 2% of all cancers are attributed to hereditary cancer syndromes^1^. Genetic risk factors for multiple primary cancers beyond known syndromes are not well understood.

Genome-wide association studies (GWAS) have implicated many common, low penetrance variants in 5p15 (*TERT-CLPTM1L*)^3^, 6p21 (*HLA*)^4,5^, 8q24^6^, and other loci in the risk of several cancer types. Additional studies have investigated pleiotropy in these regions or characterized cross-cancer susceptibility variants^7,8^. A pleiotropic locus has the potential to not only affect risk of many different cancer types, but also increase the likelihood that a single individual develops multiple primary cancers. In our prior work, we discovered that the rare pleiotropic variant *HOXB13* G84E had a stronger association with the risk of developing multiple primary cancers than of a single cancer^9^. This suggests that there may be increased power to detect pleiotropic variation in individuals with multiple primary cancers relative to those with only a single cancer. Identifying widespread pleiotropic signals is informative for understanding shared genetic mechanisms of carcinogenesis, toward the identification of informative markers for cancer prevention and precision medicine.

In this study, we survey the landscape of rare and common variation in individuals with multiple primary cancers, single cancers, and cancer-free controls through whole-exome sequencing (WES) in two large, multi-ancestry studies. We evaluate associations previously discovered in studies of individuals with a single cancer and find novel pleiotropic variation in individuals with multiple primaries.

## MATERIAL AND METHODS

### Study Populations and Phenotyping

Our study included ancestrally diverse individuals with multiple primary cancers or no cancer from two large prospective studies: the Kaiser Permanente Research Bank (KPRB) and the UK Biobank (UKB). From the KPRB, we included individuals who were previously genotyped through the Research Program on Genes, Environment and Health (RPGEH) and the ProHealth Study. For the UKB, we specifically studied participants from the 200K release of WES data, which also included individuals diagnosed with a single cancer^10^.

For both study populations, ascertainment of cancer diagnoses has been previously described^11,12^. Both studies included prevalent and incident diagnoses of malignant, borderline, and in situ primary tumors^12^. ICD codes indicating non-melanoma skin cancer or metastatic cancer were not considered primary tumors. Cancers were primarily defined according to the SEER site recode paradigm^13^. However, for hematologic cancers, we incorporated morphology following WHO classifications^14^, placing cancers into three major subtypes: lymphoid neoplasms, myeloid neoplasms, and NK- and T-cell neoplasms (Table S1). Cases were individuals with ICD-9 or ICD-10 codes for primary tumors at two or more distinct organ sites. In the KPRB, controls without a cancer diagnosis were matched 1:1 to cases on age at specimen collection, sex, genotyping array (which matched on self-reported race/ethnicity), and reagent kit. In the UKB, controls included all individuals without a cancer diagnosis.

In both study populations, we excluded duplicates/twins and first-degree relatives, retaining the individual from each related pair who had higher coverage at targeted sites. Following quality control (QC) of WES data (described below), the KPRB and UKB study populations used in this project included 3,111 and 3,318 cases with multiple primary cancers and 3,136 and 162,717 cancer-free controls, respectively. The UKB also contributed 29,091 individuals with a single cancer diagnosis. While our study was primarily unselected for cancer type, prostate cancer cases were oversampled in the KPRB due to inclusion of individuals from the ProHealth Study.

### Genetic Ancestry and Principal Components Analysis

Genetic ancestry was defined using genome-wide, imputed array data that underwent extensive QC, as previously described^12^. Ancestry principal components (PCs) were computed using flashPCA2^15^ by projecting our study samples onto PCs defined by 1000G phase 3 reference populations^16^. Individuals were assigned to the closest reference population using distance from the top 10 PCs. Individuals with ancestral PCs greater than five standard deviations from the reference population mean were excluded. The final analytic dataset included individuals of European, African, East Asian, South Asian, and Hispanic/Latino ancestry (Figure S1). A total of N = 646 (10.2%) and N= 8,739 (5.26%) individuals were of non-European ancestry in the KPRB and UKB, respectively (Table 1).

**Table 1.** Characteristics of the Kaiser Permanente Research Bank and UK Biobank study populations by ancestry group. Cases are individuals with multiple primary cancers. Controls are those without any cancer.

| Ancestry | Population: Kaiser Permanente Research Bank |  |  |  |  |  | Population: UK Biobank |  |  |  |  |  |
| --- | --- | --- | --- | --- | --- | --- | --- | --- | --- | --- | --- | --- |
|  | Cases |  |  | Controls |  |  | Cases |  |  | Controls |  |  |
|  | N | Mean Age | Female (%) | N | Mean Age | Female (%) | N | Mean Age | Female (%) | N | Mean Age | Female (%) |
| **AFR** | 99 | 70.5 | 33.3 | 100 | 70.4 | 32.0 | 29 | 55.9 | 51.7 | 3,292 | 51.8 | 60.4 |
| **EAS** | 95 | 69.7 | 49.5 | 91 | 69.5 | 49.5 | 10 | 58.8 | 80.0 | 1,009 | 52.6 | 66.9 |
| **EUR** | 2,786 | 72.8 | 43.0 | 2815 | 72.9 | 43.3 | 3,249 | 61.9 | 51.7 | 154,047 | 56.6 | 54.6 |
| **LAT** | 131 | 69.5 | 46.6 | 130 | 69.5 | 45.4 | 5 | 63.8 | 80.0 | 334 | 51.8 | 62.6 |
| **SAS** | - | - | - | - | - | - | 25 | 58.2 | 60.0 | 4,035 | 53.3 | 47.0 |

### Whole-Exome Sequencing and Quality Control

The Regeneron Genetics Center used the Illumina NovaSeq 6000 platform to perform WES for both study populations. Sample preparation and QC were performed using a high-throughput, fully-automated process that has been previously described in detail^17^. Briefly, following sequencing, reads were aligned to the GRCh38 reference genome and variants were called with WeCall^17^ for the KPRB and DeepVariant^18^ for the UKB. Samples with gender discordance, 20x coverage at less than 80% of targeted sites, and/or contamination greater than 5% were excluded.

Additional QC was applied to filter low quality variants and related individuals. First, genotype calls with low depth of coverage (DP) were updated to missing (DP < 7 for SNPs and DP < 10 for indels). Then, sites with low allele balance (AB) were removed. Specifically, variants without at least one sample having AB ≥ 15% for SNPs or AB ≥ 20% for indels were excluded. Additionally, variants with missingness > 10% and HWE p-value < 10^−15^ were excluded. Following these steps, a total of ∽3.51M high-quality sites were retained for the KPRB and ∽15.92M were retained for the UKB; excluding singletons, there were ∽1.36M and ∽8.22M variants, respectively. In the UKB, the larger number of variants observed was due to rare variation present in the larger sample size; when restricting to common variants (MAF > 1%), there were ∽186K and ∽137K variants, respectively for the KPRB and UKB.

### Association Analyses in Individuals with Multiple Cancers versus Cancer-Free Controls

Genetic association analyses of single variants and genes investigated the following cancer phenotypes: (1) diagnosis with at least two primary cancers across any of the 36 organ sites (“any 2+ primary cancers”) and (2) groupings of individuals defined by a shared index cancer at one of 16 organ sites with at least 50 cases from each study population. (“cancer-specific analyses”). Primary analyses compared multiple cancer cases to cancer-free controls. Within our cancer-specific analyses of 16 organ sites, there were cases shared across our index cancer groupings. For example, the set of individuals with at least one diagnosis of breast cancer overlaps with those having at least one ovarian cancer diagnosis.

Single-variant and gene-based association analyses were performed using REGENIE v2.2.4, a machine-learning approach for performing whole-genome regression that adjusts for case-control imbalance by applying saddlepoint approximation when the standard case-control p-value is less than 0.05^19^. We assessed single-variant associations for high-quality variants with minor allele count (MAC) > 2. WES variants were functionally annotated using SnpEff v5.0^20^ and dbNFSP v3.5^21^ accessed through ANNOVAR^22^. Missense variants were classified using five algorithms:

(1) SIFT (“D”); (2) HDIV from Polyphen2; (3) HVAR from Polyphen2; (4) LRT (“D”); and (5) MutationTaster (“A” or “D”). For our gene-based burden analyses, we used three minor allele frequency cut-offs (MAF < 0.5%, 1%, or 5%), including singletons, computed within each population. Following previous work, three gene-based models were evaluated^23^: (1) all rare variants with predicted loss-of-function (pLOF) by SnpEff, (2) pLOF and missense rare variants predicted to be deleterious by the above five classification algorithms, and (3) pLOF and missense rare variants predicted to be deleterious by at least one algorithm. Out of all allele frequency and burden combinations, we report the burden test with the lowest p-value. In the case of ties, we report the most restrictive grouping (fewest number of variants included). In our gene-based and single-variant analyses, we adjusted for covariates including age, top 10 PCs, and sex (except for sex-specific index cancers of the breast, cervix, ovary, uterus, other female genital organ, and prostate). In the KPRB population, we additionally adjusted for genotyping array and reagent kit, as they were used to perform case-control matching. In the UKB, we adjusted for flow cell (S2 vs S4), which differed for the initial 50K and subsequent 150K release of WES samples.

Single-variant and gene-based burden analyses for each phenotype were combined across study populations in a fixed-effects meta-analysis using METASOFT^24^ and metafor v3.0.2^25^, respectively. For our single-variant analyses, we report all suggestive, independent [linkage disequilibrium (LD) r^2^ < 0.2] associations with p < 5×10^−6^. For our gene-based analyses, we report all associations adjusted for the number of genes tested (p < 2.65×10^−6^ = 0.05/ 18,842). We report meta-analysis p-values (Main Text), except when a variant was unique to a single study population (Supplements).

### Distinguishing Susceptibility Signals for Multiple Cancers versus Single Cancers

We also evaluated whether the variants and genes associated with the diagnosis of multiple primary cancers (versus non-cancer controls) remained associated when comparing individuals with multiple cancers to those diagnosed with a single cancer. These analyses assessed whether the variants or genes were pleiotropic for developing multiple cancers or general markers of susceptibility to a specific cancer. We undertook these analyses in the UKB sample only, since individuals diagnosed with a single primary cancer were not sequenced in the KPRB. Single-variant and gene-level analyses were implemented as described above. For each variant or gene of interest identified in our case-control analyses, we performed a case-case analysis comparing individuals diagnosed with multiple cancers to those diagnosed with a single cancer. For our cancer-specific analyses, we compared individuals diagnosed with the index cancer plus any other cancer to those diagnosed with the index cancer only. For example, for a finding discovered in our cancer-specific analysis of prostate cancer, we performed a case-case analysis comparing individuals diagnosed with prostate cancer plus any other cancer to individuals with only a prostate cancer diagnosis.

## RESULTS

### Characterization of Multiple Primary Cancer Diagnoses in Two Large Study Populations

Our meta-analyses included 6,429 cases with multiple primary cancers and 165,853 cancer-free controls (Table 1). All cases had at least two independent primary cancer diagnoses, and 656 cases had more than two diagnoses (Figure S2). In the KPRB, the maximum number of cancer diagnoses for an individual was 6 (n = 1) and in the UKB, the maximum number was 5 (n = 2). Overall, 36 unique cancer sites were represented across multiple cancer cases in the two study populations, with 180 unique pairs of sites (e.g., breast and melanoma) and 298 unique pairs of sites and diagnostic sequence (e.g., breast followed by melanoma) (Table S2). Only 51 of the 298 ordered pairs had at least 25 cancer cases when grouping individuals by first and second cancer diagnosis (i.e., ignoring any subsequent cancer diagnoses; Table S2, Figure 1). The top ordered pairs represented in the combined study populations were prostate then melanoma (N = 221), cervix then breast (N = 202), melanoma then prostate (N = 180), breast then melanoma (N= 174), and prostate then colorectal (N = 170). Prostate, breast, melanoma, colorectal, and cervix were the most common sites of first cancer diagnoses (Figure 1). The prevalence of each cancer pair was similar in the KPRB and UKB (Figure S3). As most individual cancer pairs were underpowered for downstream analysis, we considered all multi-cancer cases combined, as well as groupings of individuals with a shared index cancer (16 cancers) (Figure S4, Table S3). Among those with multiple cancers, the cancers with the largest number of cases were prostate (N = 1,977; oversampled in KPRB), breast (N = 1,874), melanoma (N = 1,443), colorectal (N = 1,324), and urinary bladder (N = 829).

**Figure 1.**
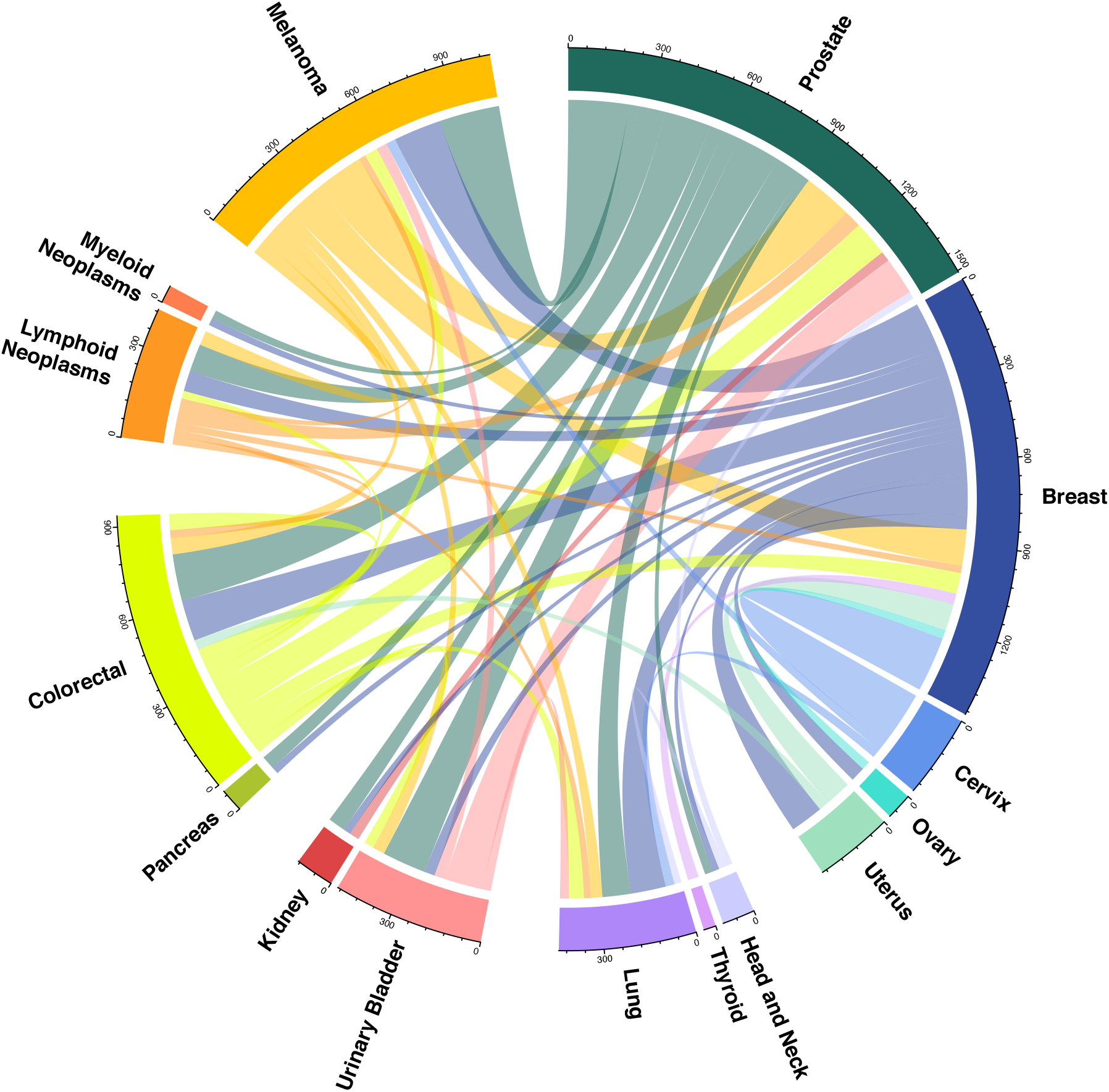
Cancer Diagnosis Pairs Present in the Combined Study Populations Circos plot describing the pairs of first and second cancer diagnoses with at least 25 cases present in Kaiser Permanente Research Bank and the UK Biobank study populations combined. Each connection reflects the number of cases with both of the linked primary cancers, where the color of the line shows the first cancer site diagnosed.

### Exome-wide Single Variant Association Analyses

We found 22 associations (p < 5×10^−6^) between individual variants and the multiple cancer phenotypes (i.e., either any 2+ primary cancers or cancer-specific analyses) (Figure 2, Table S4). We found an additional four associations (Figure S5) in our cancer-specific analyses of lymphoid and myeloid neoplasms; however, we assumed them to represent somatic alterations in the blood as they had low allele balance across our heterogenous samples (Figure S6) and occur in genes known to be impacted by clonal hematopoiesis of indeterminate potential (CHIP)^26^. Results were relatively homogeneous across the KPRB and UKB study populations (Table S4).

**Figure 2.**
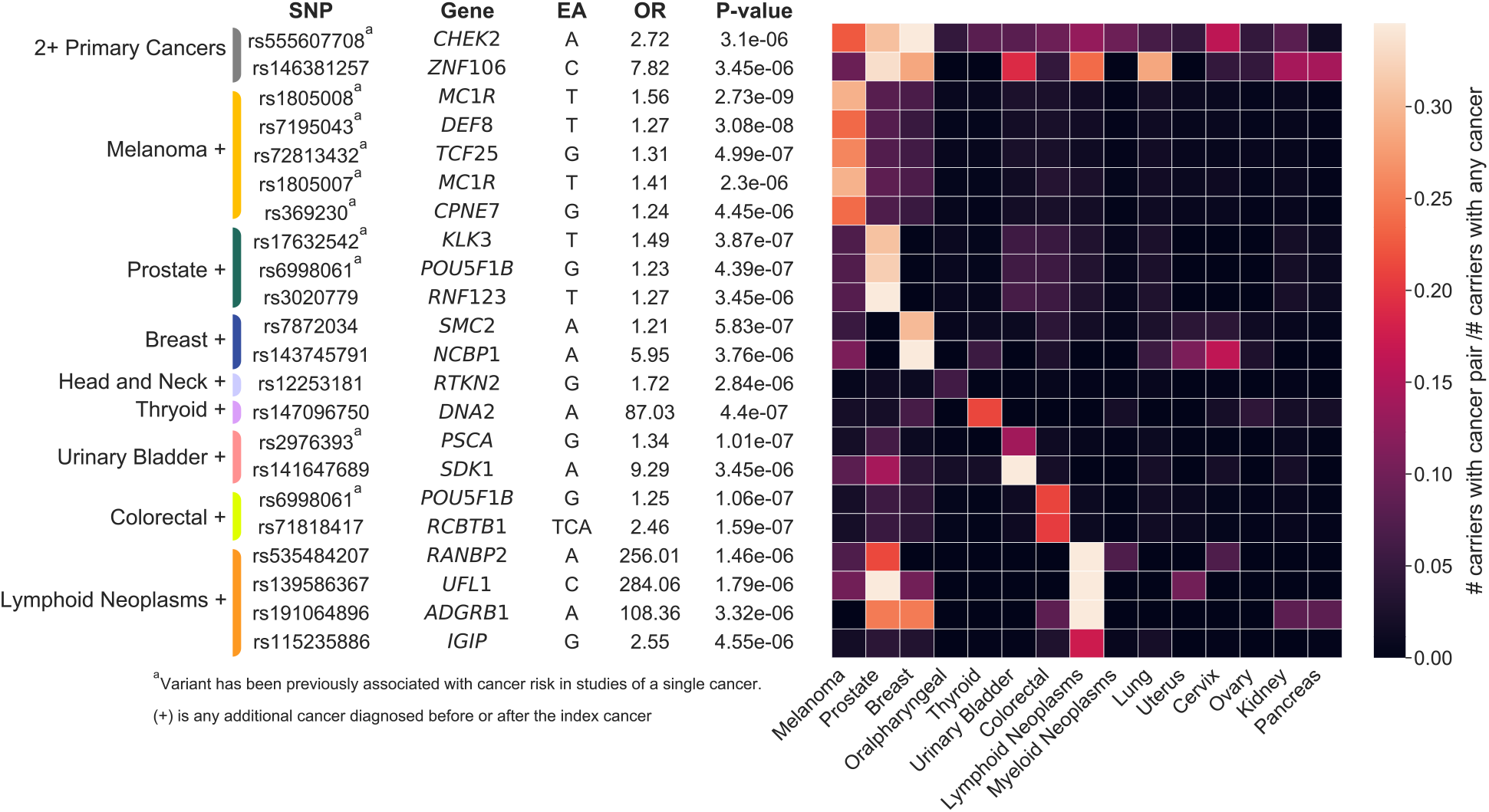
Germline Single Variant Association Results for Multiple Primary Cancers Combined or Grouped by Organ Site Suggestive (p < 5×10^−6^) germline variant associations with multiple cancer phenotypes versus cancer-free controls (n = 165,853) following a fixed-effects meta-analysis of Kaiser Permanente Research Bank and UK Biobank WES data. Associations were detected for any 2+ primary cancers (n = 6,429) and with groups of cases defined by a shared index cancer, at any time point, plus any other cancer diagnosis: melanoma + (n = 1,443), prostate + (n = 1,977), breast + (n = 1,874), head and neck + (n = 283), thyroid + (n = 198), urinary bladder + (n = 829), colorectal + (n = 1,324), lymphoid neoplasms + (n = 728). Variants that have been previously associated in single cancer studies have superscript (a). The heatmap reflects the number of carriers with the risk-increasing allele for each associated variant with the index (y-axis) and additional (x-axis) cancer over the total number of carriers, restricting to cancer cases. When the index and additional cancer are the same, the heatmap value represents all carriers with the specified cancer diagnosis divided by the total number of carriers. Abbreviations: SNP – single nucleotide polymorphism; EA – effect allele; OR – odds ratio.

We detected two variants associated with any 2+ primary cancers, rs555607708 (OR [95% CI] = 2.72 [1.79, 4.15], p = 3.10×10^−6^), a frameshift variant in *CHEK2* known to be associated with risk at many cancer sites^27^, and rs146381257 (OR [95% CI] = 7.82 [3.28, 18.62], p = 3.45×10^−6^), a 5’upstream variant in *ZNF106*. The risk-increasing allele for rs555607708 (*CHEK2*) was most commonly found among individuals with at least one breast cancer (41.9%), prostate cancer (30.6%), melanoma (22.6%), or cervical cancer (16.1%) (Figure 2). For rs146381257 (*ZNF106*), frequencies were increased in prostate cancer (33.3%), lung cancer (28.6%), breast cancer (28.6%), lymphoid neoplasms (23.8%), urinary bladder cancer (19.0%), pancreatic cancer (14.3%), and kidney cancer (14.3%).

Cancer-specific analyses identified 10 associations between previously reported risk variants for a single cancer and risk of diagnosis with that cancer plus any other cancer (Figure 2). Notably, we detected an association with the *MC1R* variant rs1805008 for melanoma^28^ (OR [95% CI] = 1.56 [1.35, 1.81], p = 2.73×10^−9^), when comparing all individuals with at least one melanoma diagnosis plus any other cancer diagnosis to cancer-free controls. We also replicated the previously associated prostate-specific antigen (PSA) variant, rs17632542^29^ (*KLK3*, OR [95% CI] = 1.49 [1.28, 1.73], p = 3.87×10^−7^) in individuals with at least one prostate cancer diagnosis. In addition, we replicated associations between missense risk variant rs6998061 (8q24 locus, *POU5F1B*) and multiple tumor types in both our prostate cancer-specific analysis^30^ (OR [95% CI]= 1.23 [1.13, 1.33], p = 4.39×10^−7^) and our colorectal cancer-specific analysis^31^ (OR [95% CI] = 1.25 [1.15, 1.37], p = 1.06×10^−7^).

The remaining variants demonstrating associations with multiple cancer phenotypes were not previously associated with any single cancer (Figure 2). They included a variant discovered in our breast cancer-specific analysis, rs143745791 (*NCBP1*, OR [95% CI] = 5.95 [2.79, 12.67], p = 3.76×10^−6^), for which 16.2% of carriers, restricted to cases, had a breast and cervical cancer diagnosis, and a variant discovered in our urinary bladder cancer-specific analysis, rs141647689 (*SDK1*, OR [95% CI] = 9.29 [3.63, 23.80], p = 3.45×10^−6^), for which 14.3% of carriers also had prostate cancer (Figure 2). Three variants found in our lymphoid neoplasm-specific analysis had increased frequencies in cases who also had a diagnosis of prostate cancer: rs535484207 (*RANBP2*, OR [95% CI] = 256.01 [26.82, 2,442.95], p = 1.46×10^−6^), rs139586367 (*UFL1*, OR [95% CI] = 284.06 [27.95, 2,886.15], p = 1.79×10^−6^), and rs191064896 (*ADGRB1*, OR [95% CI] = 108.36[15.02, 781.08], p = 3.32×10^−6^), where 21.4%, 40.0%, and 25.0% of carriers for the risk-increasing allele, for each respective variant, had both cancers. The *ADGRB1* variant was also present at increased frequencies among individuals with a lymphoid neoplasm and breast cancer diagnosis (25.0%, Figure 2).

### Gene-Based Analyses of Multiple Cancers

Out of 18,842 genes tested, we found 11 significant associations (p < 2.65×10^−6^) across our analyses of any 2+ primary cancers and our cancer-specific analyses (Figure 3, Table S5). An additional four CHIP genes (*ASXL1, TET2, JAK2*, and *DDX41*) were significantly associated with myeloid neoplasms and are likely driven by somatic alterations (Figure S7).

**Figure 3.**
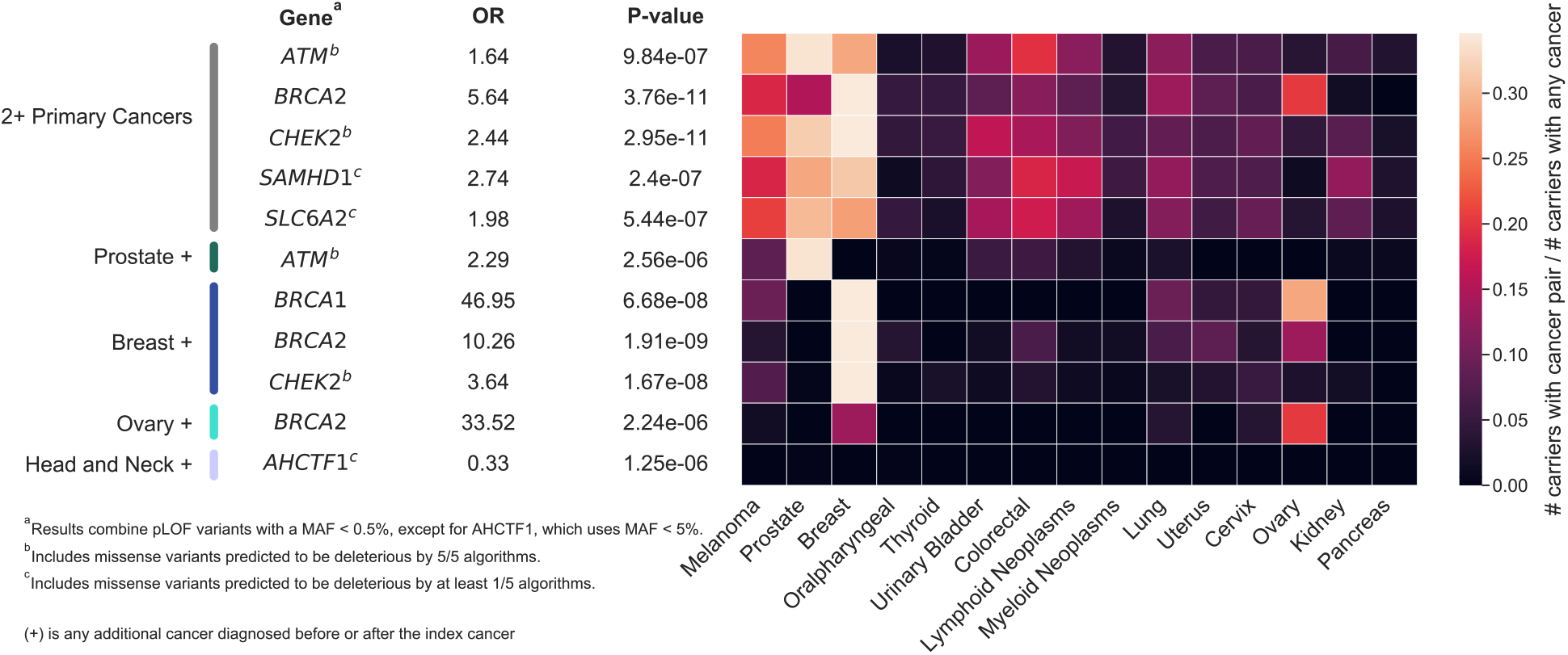
Germline Gene Based Association Results for Multiple Primary Cancers Combined or Grouped by Organ Site Burden tests were performed combining variants defined as pLOF with or without deleterious missense variants, defining deleteriousness by at least one (1/5) or all five (5/5) prediction algorithms used (Methods), at a MAF < 0.5%, 1%, or 5%. Following a fixed-effects meta-analysis of Kaiser Permanente Research Bank and UK Biobank data, Bonferroni significant associations (p < 2.65×10^−6^ = 0.05/ 18,842) corrected for the number of genes tested were found for comparisons of cancer-free controls (n = 165,853) with all cases with any 2+ primary cancers (n = 6,429) and with groups of cases defined by an index cancer for the following phenotypes: prostate + (n = 1,977), breast + (n = 1,874), ovary + (n = 239), and head and neck + (n = 283). For each gene, the variant grouping with the smallest p-value and fewest number of variants was selected. The heatmap reflects the number of carriers of each associated variant, with the index (y-axis) and additional (x-axis) cancer over the total number of carriers, where carrier is defined as having at least one alternate allele across all variants in a given gene, restricting to cancer cases. When the index and additional cancer are the same, the heatmap value represents all carriers with the specified cancer diagnosis divided by the total number of carriers. Abbreviations: OR – odds ratio; pLOF – predicted loss of function.

In our analyses of any 2+ primary cancers and our breast cancer-specific analysis, we replicated associations for known pleiotropic genes, *BRCA2* (pLOF, p = 3.76×10^−11^ and 1.91×10^−9^) and *CHEK2* (pLOF + missense, p = 2.95×10^−11^ and 1.67×10^−8^) (Figure 3). *BRCA2* also emerged in our ovarian cancer-specific analysis (pLOF, p = 1.91×10^−9^). We found associations between the known prostate cancer gene *ATM* and any 2+ primary cancers and in our prostate cancer-specific analysis (pLOF + missense, p = 9.84×10^−7^ and 2.56×10^−6^). Additional associations were observed between *SAMHD1* and *SLC642* and any 2+ primary cancers (pLOF + missense, p = 2.40×10^−7^ and p = 5.44×10^−7^, respectively). *BRCA1* also surfaced in the breast cancer-specific analysis (pLOF, p = 6.68×10^−8^), as did *AHCTF1* in the head and neck cancer-specific analysis (pLOF + missense, p = 1.25×10-6).

Functional variants in *BRCA1* and *BRCA2* were present at increased frequencies in individuals with a breast cancer diagnosis and ovary as an additional cancer site (Figure 3), such that 28.6% and 13.6% of individuals, respectively, were a carrier for at least one variant in the burden set. For *BRCA1*, there was also an increase of carriers with an additional melanoma (9.52%) or lung cancer (9.52%) diagnosis. For *BRCA2*, there was an increase of carriers with an additional uterine (8.47%), lung (6.78%), or colorectal cancer (6.78%).

### Comparison of Mutation Burden in Individuals with Multiple versus Single Cancers

Out of the 22 associated variants (above), 10 remained associated when comparing individuals with multiple cancers to those with single cancers (Table S6; p < 0.05). Two of these variants were positively associated in our analysis of any 2+ primary cancers: rs555607708 (*CHEK2*; OR [95% CI] = 1.57 [1.09, 2.25], p = 0.015) and rs146381257 (*ZNF106;* OR [95% CI] = 5.38 [1.07, 27.18], p = 0.042). The other eight variants were positively associated with diagnosis of a specific index cancer plus any other cancer versus the specific cancer alone (Table S6). Two of these eight variants were associated in our breast cancer-specific case-case analysis: rs7872034, a missense variant in *SMC2* (OR [95% CI] = 1.16 [1.05, 1.27], p = 0.0025) and rs143745791, a missense variant in *NCBP1* (OR [95% CI] = 3.71 [2.08, 6.61], p = 8.37×10^−6^).

Of the 11 findings from the gene-level burden analyses (above), seven remained positively associated with multiple cancers in comparison with single cancers (p<0.05; Table S7). Four of these genes were discovered in our case-case analysis of any 2+ primary cancers: *ATM* (OR [95% CI] = 1.20 [1.06, 1.36], p = 0.00399), *CHEK2* (OR [95% CI] = 1.56 [1.23, 1.98], p = 2.31×10^−4^), *SAMHD1* (OR [95% CI] = 1.56 [1.14, 2.13], p = 5.34×10^−3^), and *BRCA2* (OR [95% CI] = 1.86[1.31, 2.65], p = 5.43×10^−4^). *ATM* (OR [95% CI] = 1.31[1.01, 1.68], p = 0.038) was positively associated in our prostate cancer-specific case-case analysis, and the two remaining genes were positively associated in our breast cancer-specific case-case analysis: *BRCA1* (OR [95% CI] = 2.38 [1.07, 5.30], p = 0.034) and *BRCA2* (OR [95% CI] = 1.97 [1.22, 3.18], p = 0.0055).

## DISCUSSION

We investigated the genetic basis of carcinogenic pleiotropy through whole exome sequencing of individuals diagnosed with multiple primary cancers from two large, multi-ancestry study populations. Comparing individuals with multiple cancers to cancer-free controls uncovered 22 independently associated variants, ten of which remained associated when comparing individuals with multiple cancers to those with a single cancer. We also found significant associations between the genes *AHCTF1, ATM, BRCA1/2, CHEK2, SAMHD1*, and *SLC6A2* and our multiple cancer phenotypes. Other than *AHCTF1* and *SLC6A2*, these genes remained associated with multiple cancer diagnoses when comparing to individuals with a single cancer. These findings offer insights into germline exome variants that increase an individual’s risk of developing multiple primary cancers.

Compelling findings from our analyses of all individuals with more than one cancer diagnosis include associations with the rare variant rs146381257 in *ZNF106*. Carriers of the rs146381257 risk allele (C) were primarily over-represented in individuals with at least one prostate, breast, lung, or urinary bladder cancer and in individuals with lymphoid neoplasms. Carriers also demonstrated an increased risk of developing multiple cancers compared to individuals with a single cancer. *ZNF106* is an RNA binding protein involved in post-transcriptional regulation and insulin receptor signaling. Although germline variation in *ZNF106* has not previously been associated with cancer risk, a recent study found it to be associated with worse urinary bladder cancer survival^32^.

Additional noteworthy findings from our analyses of all multiple primary cancers combined include cancer susceptibility signals in *SAMHD1* and *SLC6A2*. Carriers of rare and potentially deleterious variants in *SAMHD1*, a gene with a plausible tumor suppressor role^33^, had a significantly higher risk being diagnosed with multiple cancers compared to single cancers. Germline *SAMHD1* mutations are implicated in Aicardi-Goutieres Syndrome (AGS)^34^, an autosomal recessive condition that results in autoimmune inflammatory encephalopathy. Most cancer-related studies have focused on the role of somatic alternations in *SAMHD1*^35^. However, a study of chronic lymphoid leukemia (CLL) proposed an oncogenic role of germline *SAMHD1* variation mediated by DNA repair mechanisms^36^. Consistent with this hypothesis, we also found increased *SAMHD1* variation in individuals with lymphoid neoplasms, as well as with prostate, breast, colorectal and lung cancers. *SLC6A2*, also known as *NAT1*, has been found to be prognostic for colon cancer^37^, and both in-vivo and in-vitro studies have linked expression to survival in many cancer types, including prostate^38^ and breast^39^. Polymorphisms in *SLC6A2* may also interact with smoking exposure to modulate risk for tobacco-related cancers^40^. In our study, the increased cancer risk detected among *SLC6A2* carriers was limited to comparisons with cancer-free controls.

Because we compared multiple primary cancers with both cancer-free controls and individuals diagnosed with a single cancer, we were well positioned to explore patterns of pleiotropy and disentangle variation likely to be driven by single cancers. For example, we identified two variants, rs7872034 (missense variant in *SMC2*) and rs143745791 (missense variant in *NCBP1*), associated with a diagnosis of at least one breast cancer (plus any other cancer) versus no cancer. These variants remained associated with a diagnosis of breast and another cancer when comparing to individuals diagnosed with a single breast cancer. While rs7872034 is in high LD (r2 = 0.98) with a known breast cancer risk variant (rs4742903; *SMC2* intron)^41^, it may also increase the risk of developing multiple cancers. Regarding rs143745791, germline variants in *NCBP1* have not been previously associated with cancer; because it is rare (MAF < 0.2%), larger sequencing efforts may be necessary identify variation in studies of individuals with a single cancer. Expression of this gene has been found to promote lung cancer growth and poor prognosis^42^, and *NCBP1* is overexpressed in basal-like and triple-negative breast cancers^43^. Similarly, *BRCA1/2* germline variants are prevalent among these subtypes; however, in our study populations, *BRCA1/2* carriers were more common among those with an additional ovarian cancer whereas *NCBP1* carriers more frequently had an additional cervical cancer.

In our prostate cancer-specific analysis comparing individuals with multiple cancers versus those with only a single cancer, we discovered an association with rs3020779, an eQTL for *RNF123* (also known as *KPC1*), which is a gene involved in p50 mediation and downstream stimulation of multiple tumor suppressors^44^. In our analysis of head and neck cancer, we detected an association with rs12253181 (eQTL for *RTKN2*); while this gene has not previously been associated with head and neck cancer risk, it has been shown to function as an oncogene in non-small cell lung cancer (NSCLC) and decreasing its expression may inhibit proliferation by inducing apoptosis^45^.

Limitations of our study included the identification of variants that were likely-somatic in our analyses of hematologic cancers due to an expansion of hematopoietic clonal populations with the same acquired mutation (i.e., CHIP). Confounding of germline testing by CHIP has been reported in *TP53*^46^ and *TET2*^47^, so careful interpretation is critical to avoid unnecessary clinical intervention. An additional limitation of our, and other, studies are obtaining accurate effects estimates for rare variants and the reliance on available annotations for inclusion into gene-based tests. Replication of rare findings in larger cohorts and optimization of functional impact annotations could lead to more precise results. Also, while our approach did not allow for formal replication, it was designed to identify signals for a largely understudied phenotype that were concordant in two populations. Finally, while all individuals with multiple cancers were included in our study regardless of genetic ancestry, non-European ancestries were underrepresented; larger, more diverse cohorts will be needed to fully explore the genetic basis of multiple cancers.

Strengths of this work include studying individuals of multiple ancestries who were largely unselected for specific cancer phenotypes. We also performed the first ever exome-wide study of genetic susceptibility to multiple primary cancers, using two large prospective study populations. Our study design allowed us to characterize variation across multiple primary cancers representing 36 unique sites, as well as to conduct cancer-specific analyses of 16 sites. Using this approach, we confirmed many known single-variant and gene-based findings, strengthening and supporting our novel results reported for individual cancers through our cancer-specific analyses.

In summary, by undertaking an exome-wide survey of common and rare variation in two large study populations, we identified several variant and gene-based associations that may increase the risk of developing multiple cancers within individuals. Our findings have potential implications for improving our understanding of the shared mechanisms of carcinogenesis. They may also enable screening strategies that prioritize individuals at risk for developing additional cancers. Furthermore, since many of the genes reported here have been considered as potential therapeutic targets in cancer, our work supports the use of germline information to help guide precision medicine. Future studies should aim to replicate our findings and undertake experiments that validate the functionality of the discovered pleiotropic variants. Combined with future research, our results have potential to inform genetic counseling, improve risk prediction for multiple cancers, and guide novel treatment and drug development.

## Data Availability

All results from this study are available from the article or Supplementary Materials. The UK Biobank cohort data is publicly available from the UK Biobank access portal at https://www.ukbiobank.ac.uk. The Kaiser Permanente Research Bank data are available on dbGAP. All remaining relevant data are available in the article, supplementary information, or from the corresponding author upon reasonable request.

## SUPPLEMENTAL DATA

Supplements_Tables.xls

## ACKNOWLEDGEMENTS

This material is based upon work supported by NIH grant R01 CA201358, RC2 AG036607, and the National Science Foundation Graduate Research Fellowship Program under Grant No. 1650113. Any opinions, findings, and conclusions or recommendations expressed in this material are those of the author(s) and do not necessarily reflect the views of the National Science Foundation. Support for study enrollment, survey administration, and biospecimen collection of Kaiser Permanente Research Bank participants was provided by the Robert Wood Johnson Foundation, the Wayne and Gladys Valley Foundation, the Ellison Medical Foundation, and Kaiser Permanente national and regional community benefit programs. Additionally, REG is supported by a Young Investigator Award from the Prostate Cancer Foundation. This research has been conducted using the UK Biobank Resource under Application Number 14015. Furthermore, the authors thank the Regeneron Genetics Center for covering the costs of whole-exome sequencing of the Kaiser Permanente Research Bank study participants.

## REGENERON GENETICS CENTER AUTHOR LIST AND CONTRIBUTION

### RGC Management and Leadership Team

Goncalo Abecasis, D.Phil., Aris Baras, M.D., Michael Cantor, M.D., Giovanni Coppola, M.D., Andrew Deubler, Aris Economides, Ph.D., Katia Karalis, Ph.D., Luca A. Lotta, M.D., Ph.D., John D. Overton, Ph.D., Jeffrey G. Reid, Ph.D., Katherine Siminovitch, M.D., Alan Shuldiner, M.D.

### Sequencing and Lab Operations

Christina Beechert, Caitlin Forsythe, M.S., Erin D. Fuller, Zhenhua Gu, M.S., Michael Lattari, Alexander Lopez, M.S., John D. Overton, Ph.D., Maria Sotiropoulos Padilla, M.S., Manasi Pradhan, M.S., Kia Manoochehri, B.S., Thomas D. Schleicher, M.S., Louis Widom, Sarah E. Wolf, M.S., Ricardo H. Ulloa, B.S.

### Clinical Informatics

Amelia Averitt, Ph.D., Nilanjana Banerjee, Ph.D., Michael Cantor, M.D., Dadong Li, Ph.D., Sameer Malhotra, M.D., Deepika Sharma, MHI, Jeffrey Staples, Ph.D.

### Genome Informatics

Xiaodong Bai, Ph.D., Suganthi Balasubramanian, Ph.D., Suying Bao, Ph.D., Boris Boutkov, Ph.D., Siying Chen, Ph.D., Gisu Eom, B.S., Lukas Habegger, Ph.D., Alicia Hawes, B.S., Shareef Khalid, Olga Krasheninina, M.S., Rouel Lanche, B.S., Adam J. Mansfield, B.A., Evan K. Maxwell, Ph.D., George Mitra, B.A., Mona Nafde, M.S., Sean O’Keeffe, Ph.D., Max Orelus, B.B.A., Razvan Panea, Ph.D., Tommy Polanco, B.A., Ayesha Rasool, M.S., Jeffrey G. Reid, Ph.D., William Salerno, Ph.D., Jeffrey C. Staples, Ph.D., Kathie Sun, Ph.D., Jiwen Xin, Ph.D.

## Analytical Genomics and Data Science

Goncalo Abecasis, D.Phil., Joshua Backman, Ph.D., Amy Damask, Ph.D., Lee Dobbyn, Ph.D., Manuel Allen Revez Ferreira, Ph.D., Arkopravo Ghosh, M.S., Christopher Gillies, Ph.D., Lauren Gurski, B.S., Eric Jorgenson, Ph.D., Hyun Min Kang, Ph.D., Michael Kessler, Ph.D., Jack Kosmicki, Ph.D., Alexander Li, Ph.D., Nan Lin, Ph.D., Daren Liu, M.S., Adam Locke, Ph.D., Jonathan Marchini, Ph.D., Anthony Marcketta, M.S., Joelle Mbatchou, Ph.D., Arden Moscati, Ph.D., Charles Paulding, Ph.D., Carlo Sidore, Ph.D., Eli Stahl, Ph.D., Kyoko Watanabe, Ph.D., Bin Ye, Ph.D., Blair Zhang, Ph.D., Andrey Ziyatdinov, Ph.D.

## Research Program Management & Strategic Initiatives

Marcus B. Jones, Ph.D., Jason Mighty, Ph.D., Lyndon J. Mitnaul, Ph.D.

## WEB RESOUCES

REGENIE

SnpEff

ANNOVAR

dbNSFPv3.5

flashPCA2

bcftools

plink

## DATA AVAILABILITY

All results from this study are available from the article or Supplementary Materials. The UK Biobank cohort data is publicly available from the UK Biobank access portalat https://www.ukbiobank.ac.uk. The Kaiser Permanente Research Bank data are available on dbGAP. All remaining relevant data are available in the article, supplementary information, or from the corresponding author upon reasonable request.

