## Supplemental Figures for "Assessment of Genetic Susceptibility to Multiple Primary Cancers through Whole-Exome Sequencing in Two Large Multi-Ancestry Studies"

**SUPPLEMENTAL MATERIALS**

**Figure S1.** Genetic Ancestry in the Kaiser Permanente Research Bank and UK Biobank

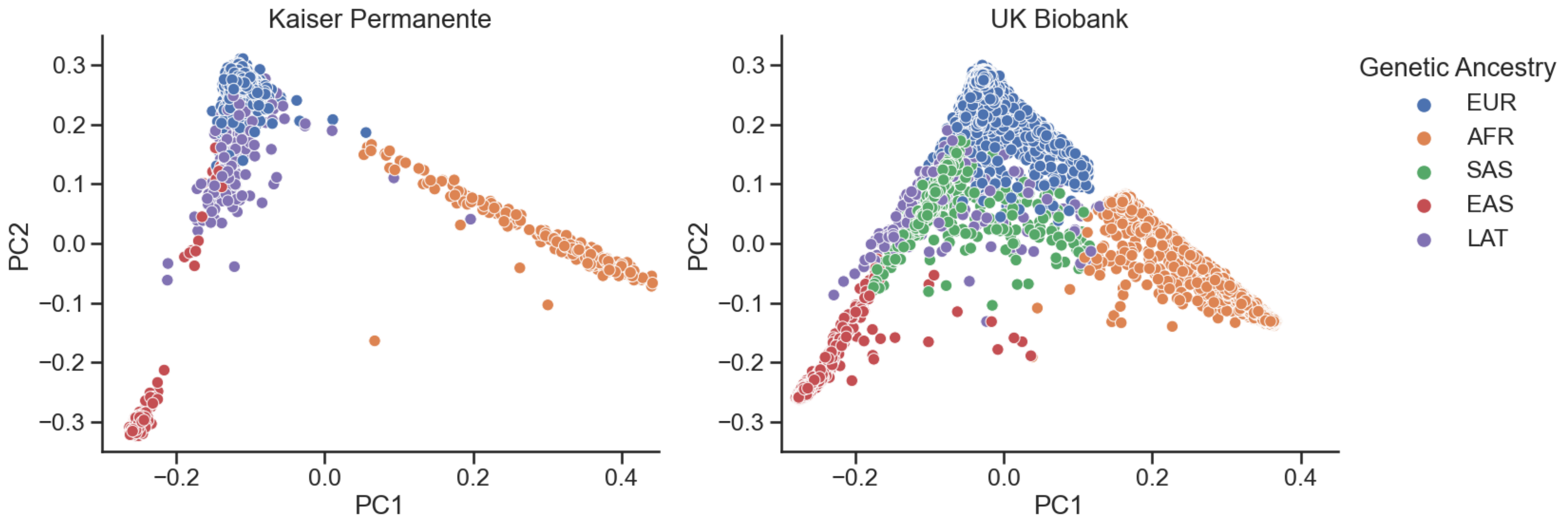


**Figure S1 Legend:** Principal components (PCs) were computed using imputed and quality-controlled genotype array data with flashPCA2. To enable accurate ancestry assignment, PCs were projected onto the 1000G phase 3 reference populations. Individuals were assigned to genetic ancestry populations using the minimum distance to the top 10 PCs, and outliers with PCs greater than 5 standard deviations from the assigned population mean were removed. Abbreviations: EUR – European; AFR – African; SAS – South Asian; EAS – East Asian; LAT – Hispanic/Latino.

**Figure S2.** Number of Primary Cancer Diagnoses and Time Intervals Between Cancer Diagnoses for Kaiser Permanente Research Bank and UK Biobank Individuals with Multiple Cancers

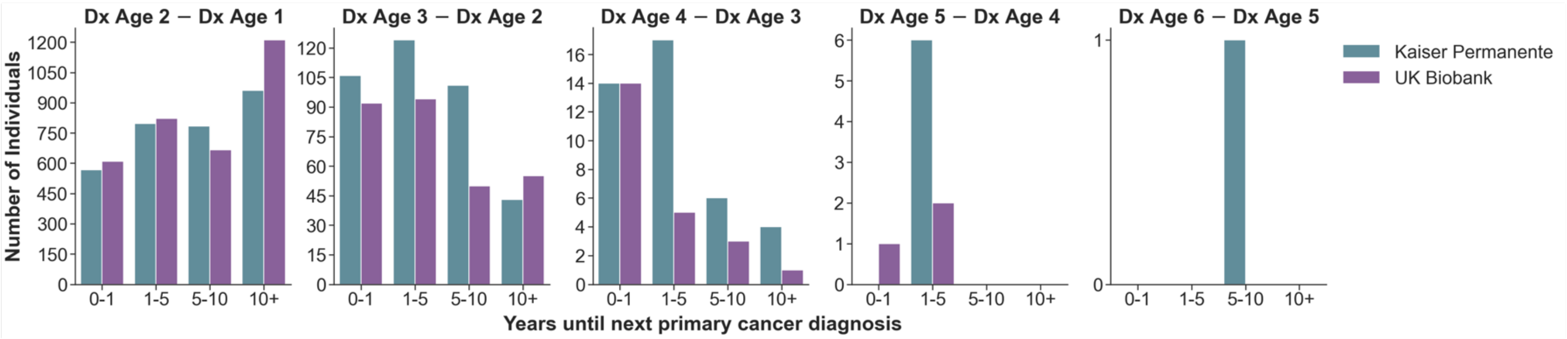


| **Number of Cancers** | **2** | **3** | **4** | **5** | **6** |
|---|---|---|---|---|---|
| **Number of Individuals** | | | | | |
| Kaiser | 2,737 | 333 | 35 | 5 | 1 |
| UK Biobank | 3,020 | 268 | 20 | 3 | 0 |

**Figure S2 Legend:** The number of primary cancer diagnoses for each individual was tabulated for the Kaiser Permanente Research Bank and UK Biobank study populations. The above bar plot shows individuals binned according to time between cancer diagnoses, reflecting the subsequent diagnosis occurring less than 1 year, 1 to 5 years, 5 to 10 years, or more than 10 years after the prior diagnosis. Abbreviations: Dx – Diagnosis

**Figure S3.** Most Common Cancer Pairs Present in Kaiser Permanente and UK Biobank Cases with Multiple Cancers

**Population: Kaiser Permanente**

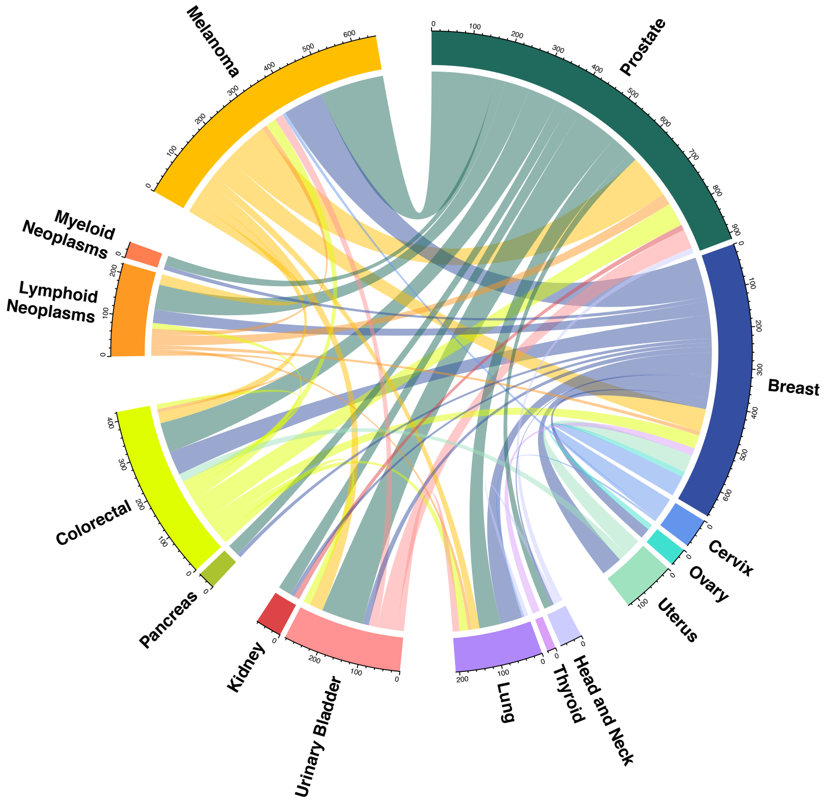


**Population: UK Biobank**

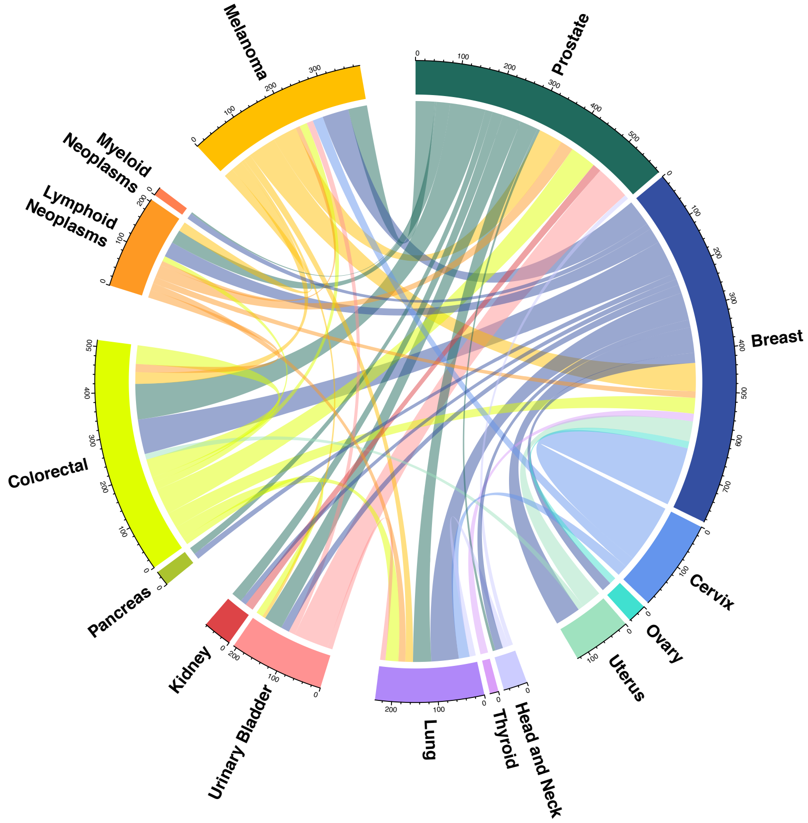


**Figure S3 Legend:** Circos plots describing pairs of first and second cancer diagnoses with at least 25 cases present in the Kaiser Permanente Research Bank (left) and UK Biobank (right). Each connection reflects the number of individuals with both of the linked primary cancers, where the color of the line shows the first cancer site diagnosed.

**Figure S4.** Cancers Represented in the Kaiser Permanente Research Bank and UK Biobank with Sufficient Sample Size for Exome-wide Association Analyses

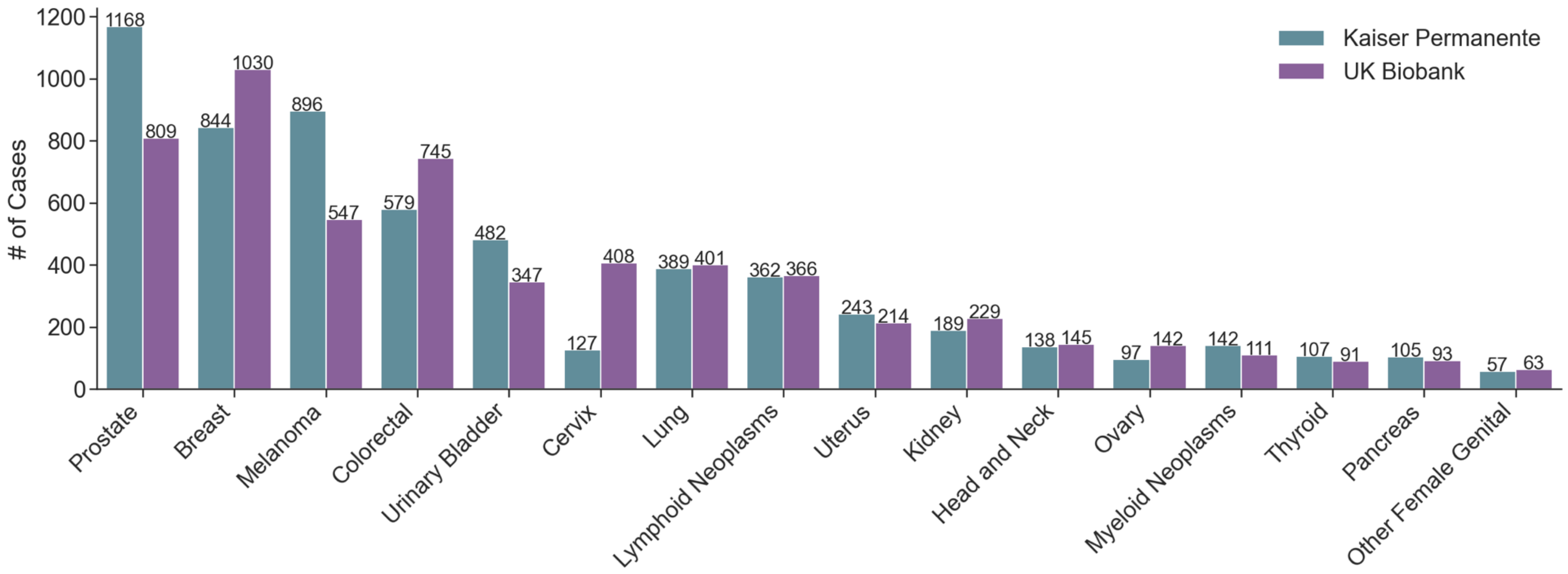


**Figure S4 Legend:** All individuals with multiple cancers who were diagnosed with cancer at a specific site, at any time point, were grouped together. A total of 16 cancer sites (represented above) had sufficient sample size (N > 50) in each study population for downstream association analyses.

**Figure S5.** Significant Single-Variant Association Results Due to Clonal Hematopoiesis of Indeterminate Potential

| | SNP | Gene | EA | OR | P-value |
|---|---|---|---|---|---|
| Lymphoid Neoplasms + | rs188201162 | *PDE4DIP* | T | 3.14 | 4.62e-06 |
| Myeloid Neoplasms + | rs756958159 | *ASXL1* | AG | 183.54 | 1.46e-11 |
| | rs751713049 | *SRSF2* | T | 1126.32 | 6.66e-10 |
| | rs559063155 | *SF3B1* | C | 2301.65 | 2.38e-06 |

(+) is any additional cancer diagnosed before or after the index cancer

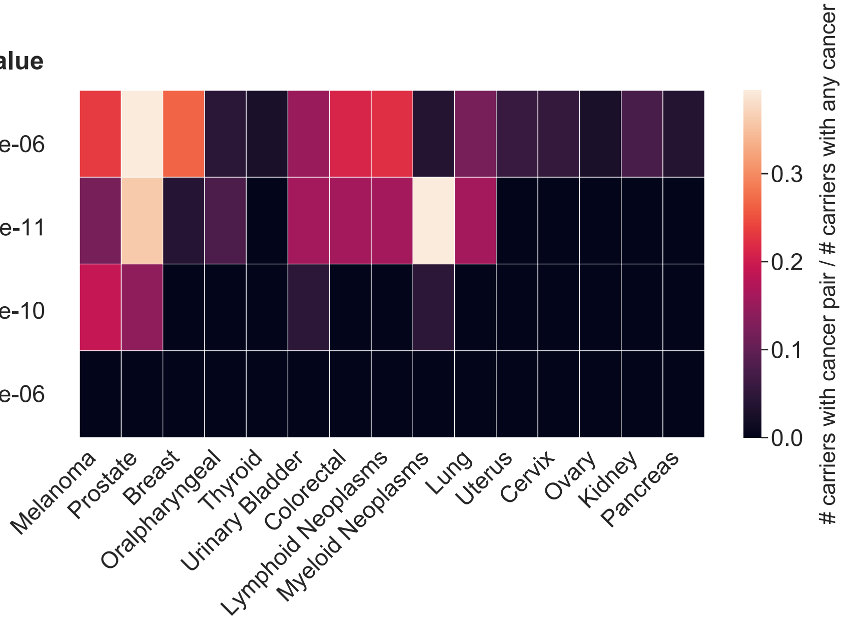


**Figure S5 Legend:** Significant variant associations ($p < 5x10^{-6}$) with blood cancer phenotypes compared to cancer-free controls following a fixed-effects meta-analysis of the Kaiser Permanente Research Bank and UK Biobank WES data. Because of low allele balance, which is suggestive of confounding by clonal hematopoiesis of indeterminate potential (CHIP), mutations are expected to be somatic. The heatmap reflects the number of carriers with the risk-increasing allele for each associated variant with the index (y-axis) and additional (x-axis) cancer over the total number of carriers. When the index and additional cancer are the same, the heatmap value represents all carriers with the specified cancer diagnosis divided by the total number of carriers, restricting to cancer cases.

**Figure S6.** Allele Balance for Findings Related to Lymphoid and Myeloid Neoplasms

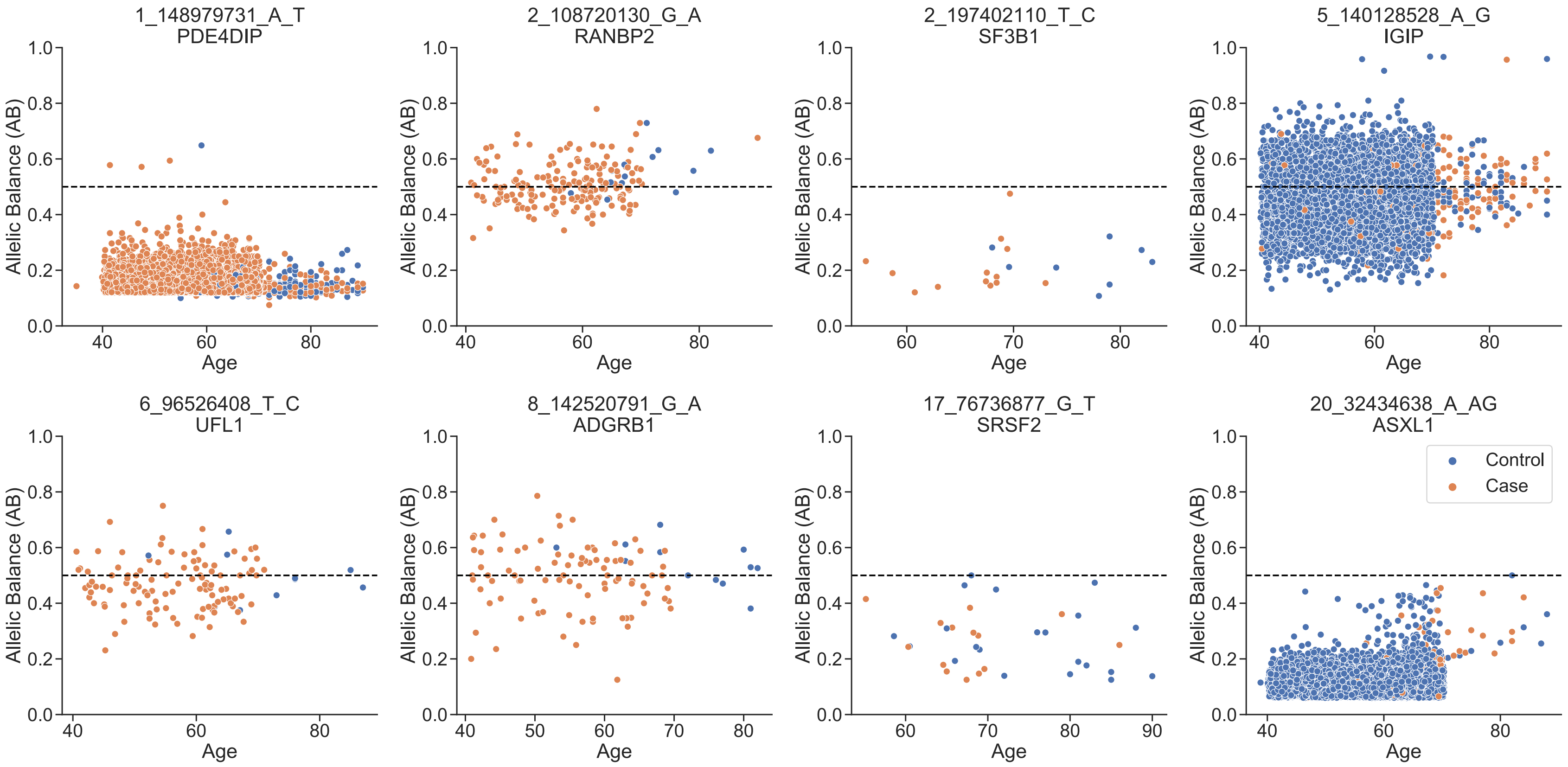


**Figure S6 Legend:** For significant variants discovered in our cancer-specific analyses of either myeloid or lymphoid neoplasms, we looked at the allele balance as a function of age for individuals heterozygous at each locus. If the majority of heterozygous individuals had allele balance below 0.5, we assumed that they were likely somatic due to confounding by clonal hematopoiesis of indeterminate potential (CHIP).

**Figure S7.** Significant Gene-Based Association Results Due to Clonal Hematopoiesis of Indeterminate Potential

| | Gene[a] | OR | P-value |
|---|---|---|---|
| Myeloid Neoplasms + | *ASXL1* | 88.59 | 3.53e-13 |
| | *DDX41*[b] | 44.93 | 1.04e-06 |
| | *JAK2*[c] | 38.94 | 3.7e-13 |
| | *TET2* | 38.86 | 5.54e-10 |

[a] Results combine pLOF variants with a MAF < 0.5%, except for AHCTF1, which uses MAF < 5%.
[b] Includes missense variants predicted to be deleterious by 5/5 algorithms.
[c] Includes missense variants predicted to be deleterious by at least 1/5 algorithms.

(+) is any additional cancer diagnosed before or after the index cancer

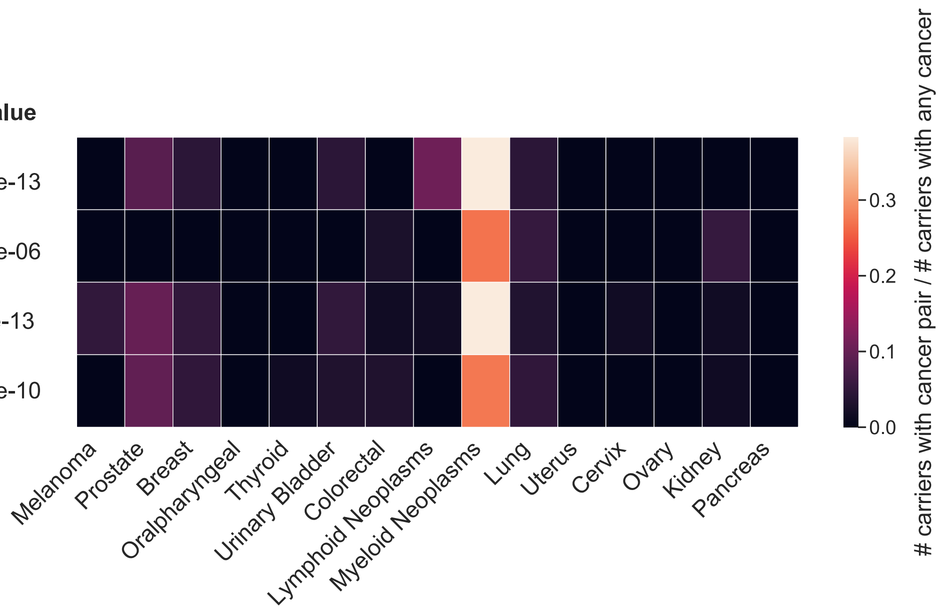


**Figure S7 Legend:** Burden tests were performed combining variants defined as pLOF with or without deleterious missense variants, defining deleteriousness by at least one (1/5) or all five (5/5) of prediction algorithms used (Methods), at a minor allele frequency < 0.5%, 1%, or 5%. Bonferroni significant associations ($P < 2.65\times10^{-6} = 0.05/ 18,842$) corrected for the number of genes tested were found for comparisons of cancer-free controls with myeloid neoplasms following a fixed-effects meta-analysis of Kaiser Permanente Research Bank and UK Biobank data. For each gene, the variant grouping with the smallest p-value and fewest number of variants was selected. The heatmap reflects the number of carriers of each associated variant, with the index (y-axis) and additional (x-axis) cancer over the total number of carriers where carrier is defined as having at least one alternate allele across all variants in a given gene. When the index and additional cancer are the same, the heatmap value represents all carriers with the specified cancer diagnosis divided by the total number of carriers, restricting to cancer cases. Myeloid associations occur in frequently mutated clonal hematopoiesis of indeterminate potential (CHIP) genes.
